# Dynamics of infection-elicited SARS-CoV-2 antibodies in children over time

**DOI:** 10.1101/2022.01.14.22269235

**Authors:** Lauren E. Gentles, Leanne Kehoe, Katharine H.D. Crawford, Kirsten Lacombe, Jane Dickerson, Caitlin Wolf, Joanna Yuan, Susanna Schuler, John T. Watson, Sankan Nyanseor, Melissa Briggs-Hagen, Sharon Saydah, Claire M. Midgley, Kimberly Pringle, Helen Chu, Jesse D. Bloom, Janet A. Englund

## Abstract

Severe acute respiratory syndrome coronavirus 2 (SARS-CoV-2) infection elicits an antibody response that targets several viral proteins including spike (S) and nucleocapsid (N); S is the major target of neutralizing antibodies. Here, we assess levels of anti-N binding antibodies and anti-S neutralizing antibodies in unvaccinated children compared with unvaccinated older adults following infection. Specifically, we examine neutralization and anti-N binding by sera collected up to 52 weeks following SARS-CoV-2 infection in children and compare these to a cohort of adults, including older adults, most of whom had mild infections that did not require hospitalization. Neutralizing antibody titers were lower in children than adults early after infection, but by 6 months titers were similar between age groups. The neutralizing activity of the children’s sera decreased modestly from one to six months; a pattern that was not significantly different from that observed in adults. However, infection of children induced much lower levels of anti-N antibodies than in adults, and levels of these anti-N antibodies decreased more rapidly in children than in adults, including older adults. These results highlight age-related differences in the antibody responses to SARS-CoV-2 proteins and, as vaccines for children are introduced, may provide comparator data for the longevity of infection-elicited and vaccination-induced neutralizing antibody responses.

## Introduction

SARS-CoV-2, the causative agent of coronavirus disease 2019 (COVID-19), elicits an antibody response targeting multiple viral proteins following infection. Anti-spike (S) antibodies are of particular importance because S is the major target of neutralizing antibodies and neutralizing anti-S antibody titers correlate with protection (1–4). For this reason, currently authorized vaccines only include the S antigen and specifically induce anti-S responses. Additionally, SARS-CoV-2 neutralization assays are designed to measure the potency of antibodies that block viral binding and entry to cells, including via inhibiting S binding to host angiotensin converting enzyme 2 (ACE2) receptor on host cells, and/or inhibiting S fusion. Nucleocapsid (N) protein is also highly immunogenic during SARS-CoV-2 infection and is a predominant target of binding antibodies making it a robust marker of infection. In adults, circulating antibodies rise to peak titers within 3-5 weeks after infection and then gradually begin to wane (1, 3, 5–14). Studies have shown a strong positive correlation between neutralizing antibody titers and protection from subsequent infection (4, 15–19).

COVID-19 in children tends to be milder than in adults, resulting in lower risk of progression to hospitalization and death (20, 21). However, clinical manifestations of COVID-19 vary widely in children as in adults and can range from asymptomatic infections to illness lasting for several months (22). Furthermore, infection by SARS-CoV-2 in children causes a greater burden of hospitalization and death than the pre-vaccine burden of some common childhood illnesses, including varicella (23). Previous work has documented the acute and convalescent dynamics of the SARS-CoV-2 antibody response in adults across a wide range of ages and disease severities (1, 3, 8, 10, 11, 14, 23, 24), but fewer data are available detailing the longevity of circulating antibodies in the pediatric population (24–27).

Here, we follow a cohort of 32 SARS-CoV-2-infected convalescent children <18 years old for up to 52 weeks post-symptom onset, measuring anti-S neutralizing antibody levels with a pseudoneutralization assay, and anti-N binding antibody levels. We compare the pediatric antibody response to those in a previously characterized cohort of adults (3).

## Materials and Methods

### Pediatric Participants

Our IRB-approved study enabled us to enroll children, defined as <18 years old at enrollment, including children with underlying medical conditions, and obtain sera for the assessment of immune responses to SARS-CoV-2 infection at Seattle Children’s Hospital, Seattle, WA, beginning in April 2020. Informed consent was obtained from parents and assent from children over 7 years of age. The REDCap electronic data collection tool was used to acquire demographics, hospitalization data; clinical information including respiratory support, ICU admission, length of stay; laboratory studies including viral testing results, and medical history including chronic underlying medical conditions (28). This study was reviewed and approved by the Seattle Children’s Hospital IRB^§^.

Children with confirmed or presumed SARS-CoV-2 infection were recruited to our study during April 2020 through January 2021. Children were considered to have a confirmed SARS-CoV-2 infection if they tested positive for SARS-CoV-2 by RT-PCR. Children were presumed to have SARS-CoV-2 infection if they did not have documentation of a positive RT-PCR, but had detectable SARS-CoV-2-specific antibodies and either: 1) presented with confirmed Multisystem Inflammatory Syndrome in Children (MIS-C), or 2) were symptomatic and had an RT-PCR-positive household contact. Reported symptoms included but were not limited to sore throat, cough, fever, loss of taste or smell, fatigue, runny nose, head ache, and diarrhea.

Enrollment included hospitalized children, children who were tested for SARS-CoV-2 using RT-PCR as outpatients as determined by their provider, and children who did not receive medical care but were recruited from the community, including community-based surveillance platforms (29). Children were recruited during acute illness with sera drawn at approximately 4-8 weeks (1-2 months), 24 weeks (6 months), and 52 weeks (12 months) following symptom onset for confirmed or presumed infection. Only children who provided at least two specimens by May 2021 were included in this analysis. In addition, only presumed cases with at least one positive serological result were included (**Supplemental Table 1**). For asymptomatic cases, weeks post-positive RT-PCR test result was used as a substitute for weeks post-symptom onset. For children who developed MIS-C, “weeks post-symptom onset” refers to acute infection symptoms before MIS-C onset. No children in this study were vaccinated prior to specimen collection.

### Adult Participants

Adult specimens were collected as a part of the Hospitalized or Ambulatory Adults with Respiratory Viral Infections (HAARVI) cohort at the University of Washington Department of Medicine (3, 30, 31). Adults were enrolled from March through May of 2020. A convenience sample of adults who provided specimens at roughly eight- and twenty-four-weeks post-symptom onset were included in this analysis. Study enrollment and specimen collection are detailed elsewhere (3, 30, 31). Briefly, adults were enrolled in the study following RT-PCR confirmed SARS-CoV-2 infection. Inpatients were recruited for enrollment during their hospital stay at Harborview Medical Hospital, University of Washington Medical Center, or Northwest Hospital in Seattle, Washington in 2020. Asymptomatic adults were identified as participants who responded “None” to a symptom questionnaire and tested positive for SARS-CoV-2 infection via outpatient or community testing. Informed consent was provided by all participants or their legally authorized representatives. No adults in this study were vaccinated prior to specimen collections since no vaccines were available during the collection period, and no adults in this study were enrolled in ongoing vaccine clinical trials. Weeks post-positive RT-PCR test result was used in lieu of weeks post-symptom onset for asymptomatic adults.

### Laboratory Methods

#### Pediatric specimen collection

Whole blood collection was scheduled for 4 to 8-weeks, 24-weeks, and 52-weeks post-symptom onset for the pediatric cohort (**Supplemental figure 1**). Blood specimens were collected in serum separator tubes, stored at 5°C, and spun within 24 hours before being aliquoted and stored at -80☐. Heat inactivation of all specimens was performed at 56☐ for 30 minutes before performing serological assays.

**Figure 1.**
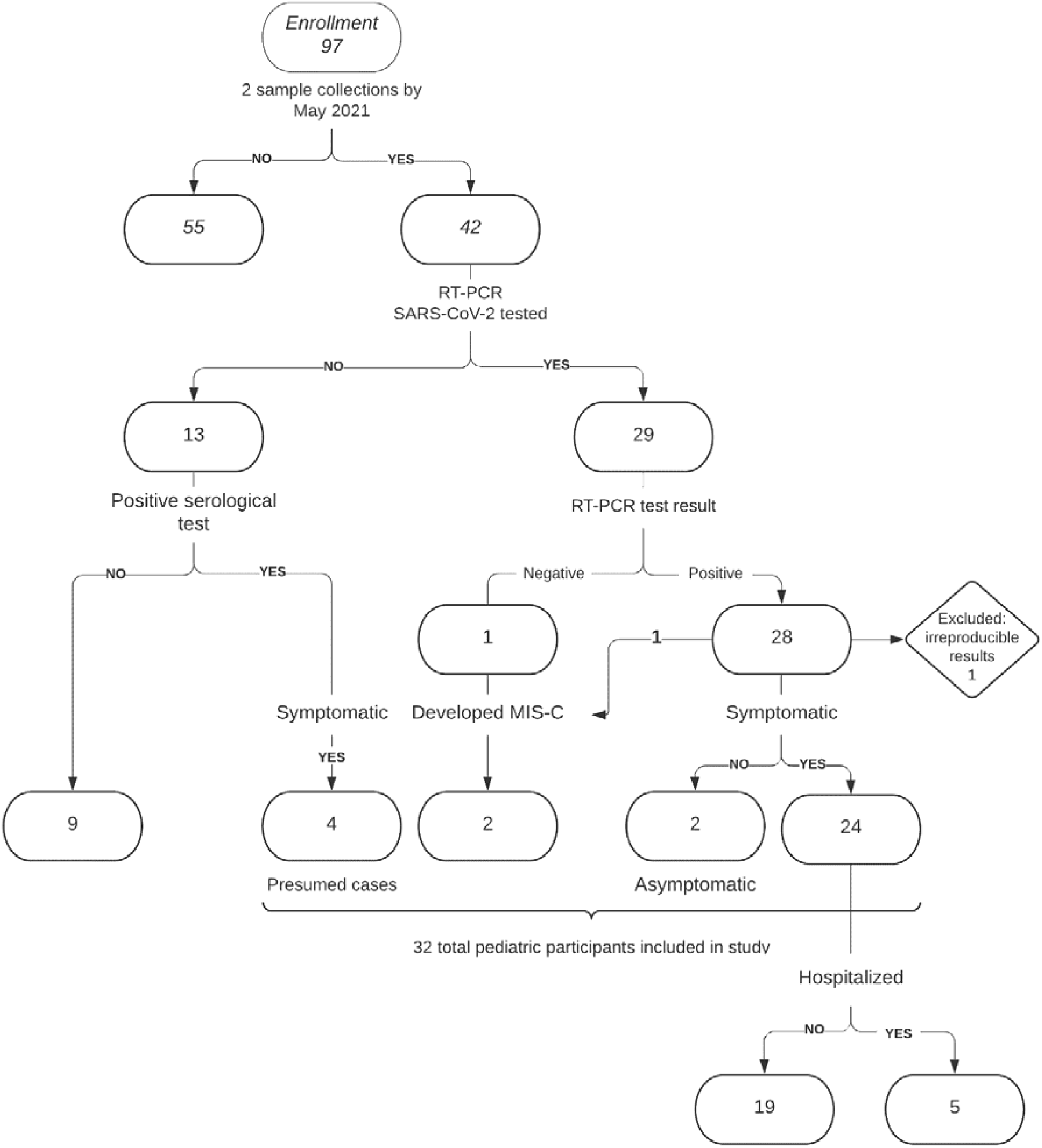
Pediatric study inclusion criteria flowchart. Evidence of infection included a PCR-positive test (n=28) or positive serological test result following a known RT-PCR-positive household exposure (n=4) and/or presentation with MIS-C (n=2).

#### Adult specimen collection

Whole blood collection was scheduled for 8- and 24-weeks post-symptom onset for the adult cohort. Blood specimens were immediately added to acid citrate dextrose tubes upon collection which were then spun down to separate out the red blood cell fraction. Within 6 hours following collection, aliquots of these specimens were frozen at -20☐ for storage. Prior to use in serological assays, all specimens were heat inactivated at 56☐for one hour.

#### Neutralization assays

Neutralization assays were performed as previously reported using spike-pseudotyped lentiviral particles (3). The spike protein used is based on Wuhan-Hu-1 (GenBank: <u>MN908947</u>) with a 21 base pair deletion (delta21) at the terminus of the cytoplasmic tail that enhances viral titers (32– 37). The spike also contains the mutation D614G that has become predominant in circulating strains (38). Plasmid HDM_Spikedelta21_D614G encoding this spike protein is available from AddGene (no. 155130) or BEI Resources (NR-53765) along with the full annotated sequence. To perform neutralization assays, 1.25x10^4^ HEK-293T-ACE2 cells (39) (BEI resources NR-52511) are added in 50ul per well of a 96-well poly-L-lysine coated plate (Greiner; no. 655936). Our limit of detection for the neutralization assay is 1:20 since this is the starting serum dilution. All assays included pre-pandemic pooled serum collected between 2015 to 2018 as a negative control. No substantial neutralization was observed for a pool of pre-pandemic sera at a dilution of 1:20. SARS2 Spike-D614G-delta21 pseudotyped lentivirus particles encoding luciferase were added at a dilution of 200,000 RLU per well as determined by titering. The virus-antibody plate was then incubated for 1 hour at 37°C before being added to the plate with cells. Neutralization titers were determined using a plate reader to measure luciferase activity at 50 hours post-infection. Measurements were given as the reciprocal dilution of sera at which viral infection was inhibited by 50% (NT_50_). NT_50_ values were calculated using the neutcurve python package version 0.5.3 available here: https://github.com/jbloomlab/neutcurve which fit a Hill curve to our data to determine the 50% inhibitory concentration (IC_50_). NT_50_ values reported here were the reciprocal of the IC_50_.

#### SARS-CoV-2 IgG assay

The SARS-CoV-2 IgG assay, an FDA Emergency Use Authorized immunoassay, which utilizes a chemiluminescent test to assess immunoglobulin G (IgG) binding to nucleocapsid (N) protein, was performed according to manufacturer specifications (Abbott). Anti-N IgG index values were assessed; higher index values reflected higher antibody levels. An index value of > 1.40 is considered a positive result for this assay. Sensitivity and specificity of the SARS-CoV-2 IgG assay have been reported elsewhere (23, 40–44).

#### Comparison of antibody levels in a subset of immunocompetent children and adults

For comparison of antibody levels between pediatric participants and adults, we limited our analysis to only specimens that were collected within a similar range of weeks post-onset between 8-13 (first collection period) and 24-29 (second collection period) weeks for both cohorts. In this sub-analysis, we excluded participants with MIS-C development, complicating immunocompromising conditions, or receipt of multiple blood transfusions. We assessed changes in antibody titers over time among a limited number of children and adults with two specimens collected within these comparative time frames. Statistical significance was determined by Mann Whitney test.

## Results

### Study participants

From April 2020 through June 2021, we enrolled 97 pediatric participants of whom 42 had completed at least 6-months of follow-up with two blood draws obtained by May 2021 (**Figure 1**). Thirty-two of the 42 children had evidence of confirmed or presumed infection and were included in the pediatric analysis: 27 of 32 had a confirmed positive RT-PCR test, including one of two children who presented with MIS-C; one of 32 had a positive serological test result and presented with MIS-C; and four of 32 had a positive serological test result and a known RT-PCR-positive household member (**Supplemental Table 1**). Among the 32 children included in this analysis, median age was 12 years, 6 (19%) were female, 5 (16%) were symptomatic and hospitalized, 25 (78%) were symptomatic but not hospitalized, and 2 (6%) were asymptomatic during acute infection (**Table 1, Figure 2**). Of the two children who developed MIS-C: one (C27) had an asymptomatic acute infection (identified through RT-PCR) and subsequently required ICU admission and supplemental oxygen in the form of bilevel positive airway pressure upon the onset of MIS-C symptoms; the other (C15) had an initial SARS-CoV-2 respiratory infection managed as an outpatient but was subsequently hospitalized with MIS-C, during which time C15 was SARS-CoV-2 RNA-negative and antibody-positive.

**Figure 2:**
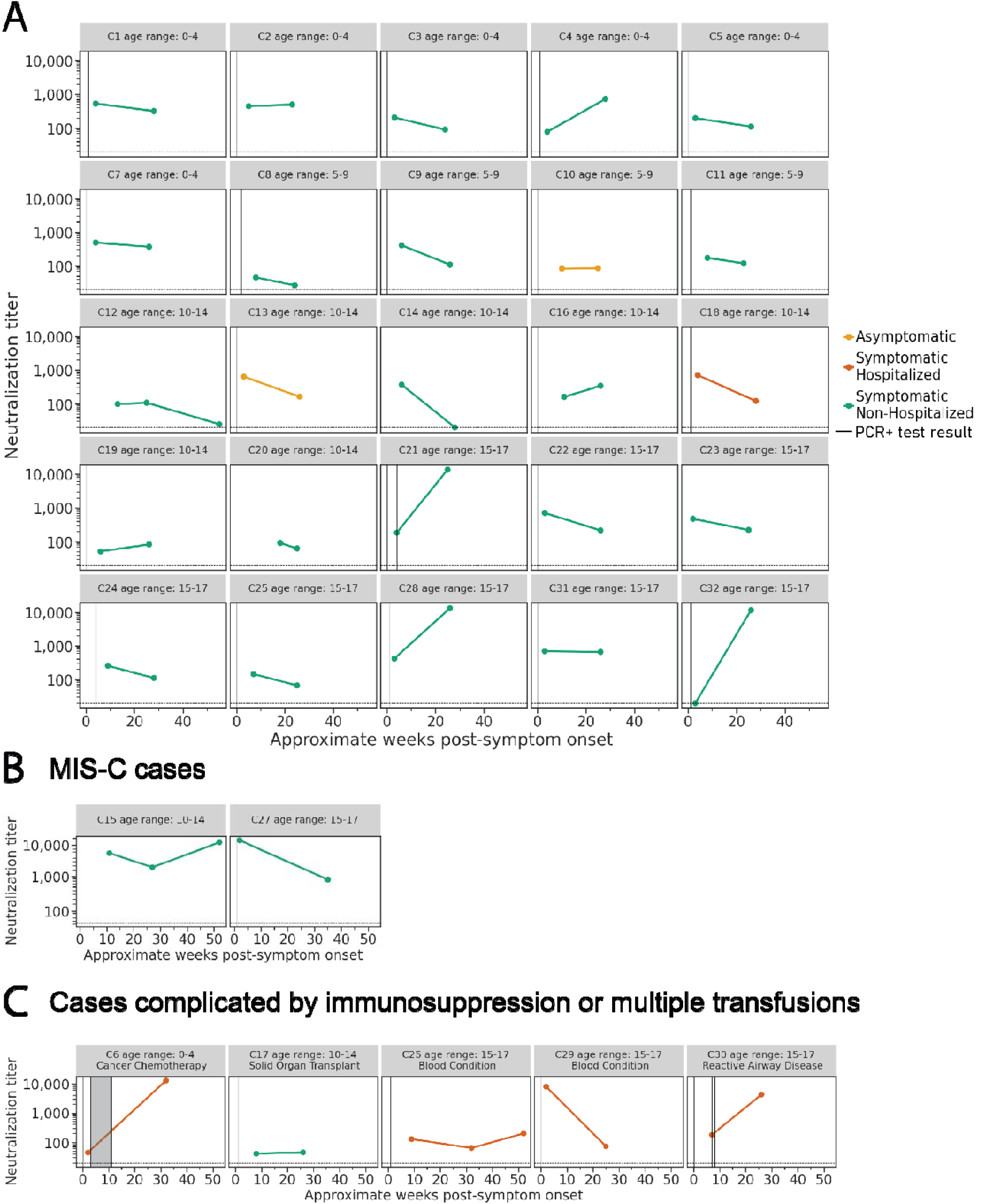
Neutralization titers in children over time. Neutralizing antibody titers (NT_50_) in **A)** 25 children with confirmed SARS-CoV-2 infection, **B)** 2 children who developed MIS-C following acute infection and **C)** cases complicated by immunosuppression (N = 4) or multiple blood transfusions (N = 1) in 5 children with confirmed SARS-CoV-2 infection followed prospectively over time shown as weeks. Vertical lines represent the week of positive RT-PCR test result(s), and shaded areas indicate weeks with consecutive positive RT-PCR test results. Colors show disease severity during acute infection. Dotted horizontal lines indicate the limit of detection (20).

**Table 1.** Pediatric and adult cohort demographics by disease severity.

**Pediatric cohort**
| Characteristic | Asymptomatic<br>n=2 | Symptomatic<br>non-hospitalized<br>n=25 | Symptomatic<br>hospitalized<br>n=5 | Overall<br>n=32 |
| --- | --- | --- | --- | --- |
| Age, median (range) | 10 (9.3-10.7) | 11.8 (0.2-17.8) | 16 (3.6-17.7) | 12 (0.2-17.8) |
| Sex, no. (%) |  |  |  |  |
| Female | 0 (0) | 4 (16) | 2 (40) | 6 (19) |
| Male | 2 (100) | 21 (84) | 3 (60) | 26 (81) |
| Immunocompromised or received multiple blood transfusions* | 0 | 1 | 4 | 5 |

\*No other children reported chronic conditions.

**Adult cohort**
| Characteristic | Asymptomatic<br>n=4 | Symptomatic<br>non-hospitalized<br>n=8 | Symptomatic<br>hospitalized<br>n=2 | Overall<br>n=14 |
| --- | --- | --- | --- | --- |
| Age, median (range) | 69.5 (60-79) | 65 (47-76) | 59 (54-64) | 65 (47-79) |
| Sex, no. (%) |  |  |  |  |
| Female | 3 (75) | 4 (50) | 1 (50) | 8 (57) |
| Male | 1 (25) | 4 (50) | 1 (50) | 6 (43) |

Five children had underlying immunocompromising conditions or received multiple blood transfusions; four of whom were hospitalized. Among the 25 children who were not immunocompromised, did not receive multiple blood transfusions, and did not present with MIS-C (**Figure 2A**), one child was hospitalized, 22 children were symptomatic but not hospitalized, and two children were asymptomatic.

A second cohort of 14 SARS-CoV-2-infected unvaccinated immunocompetent adults between the ages of 47 and 79 years (median: 65) was included in this study as a comparator group. We previously profiled neutralizing antibody dynamics for all these adults out to 90 days post-symptom onset (3) (See **Supplemental Table 2**). Here we performed additional assays for the same adult participants to enable direct comparison with the pediatric cohort in a sub-analysis. This convenience sample of 14 adults included two who were symptomatic and hospitalized, 8 who were symptomatic non-hospitalized, and 4 who were asymptomatic. Eight (57%) adults were female. Two adult participants reported underlying conditions: one participant (A3) was recorded as having diabetes, chronic obstructive pulmonary disease, asthma, and obstructive sleep apnea; and another (A13) had hypertension.

### Specimen collection

During the 4- and 24-week pediatric blood collections, specimens were collected from the 32 children at a median of 4.5 weeks (IQR: 2.5weeks; range: 2-18weeks) and 26 weeks (IQR: 1.25weeks; range: 23-35weeks), respectively; 3 children also had blood collected at 52 weeks. At 8- and 24-weeks, specimens were collected from the 14 adults at a median of 9.5 (range: 8-13weeks, IQR:1wk) and 25 weeks (range: 24-29weeks, IQR: 1wk), respectively. To compare pediatric and adult responses, we performed a sub-analysis which included specimens collected within two collection periods: the first at 8-13 weeks, and the second at 24-29 weeks. This sub-analysis included specimens from all 14 adults; for children, 7 children had blood drawn in the first collection period (median = 9.5 weeks; IQR = 2.5) and 24 children had blood drawn in the second collection period (median 26 weeks; IQR=1). Five children and 14 adults, with specimens collected at both timepoints, were included in fold-change analyses.

### Neutralization dynamics over time in children

We measured neutralization titers for the pediatric specimens collected at each time period (**Figure 2A, B, & C**). All children with confirmed or presumed infections had measurable neutralizing antibody titers for at least one specimen. For the 25 children without MIS-C or immunocompromising conditions or multiple blood transfusions, overall neutralization titers changed very little over the course of 24 weeks from a geometric mean NT_50_ of 214 and 244 for the first and second collection period, respectively. Interestingly, a greater than 4-fold increase in neutralization titer between the first and second collection period was seen for four children all of whom were symptomatic but not hospitalized. If these four children are excluded, the geometric mean NT_50_ decreases by 1.86-fold from the first to the second collection period (from 245 to 132, respectively). For two of the 25 children without immunocompromising conditions, a decrease of greater than 4-fold between 4 and 24 weeks was observed. Both children were symptomatic of whom one was hospitalized. For 19 (76%) of the 25 children, less than 4-fold (range 3.86- to 1.02-fold) changes in neutralization titers were observed. One child with increasing titers, (C32), had no detectable neutralization titer at 3 weeks post-symptom onset despite testing positive by RT-PCR, but subsequently developed high neutralization titers by 26 weeks. Despite the variability among individual immunocompetent children, some trends in the overall antibody dynamics were observed (**Figure 3A**). Nearly all immunocompetent children had neutralizing activity at all timepoints, and the majority of children (15 out of the 25 total) exhibited at least a 25% decrease in neutralization titers over 24 weeks.

**Figure 3.**
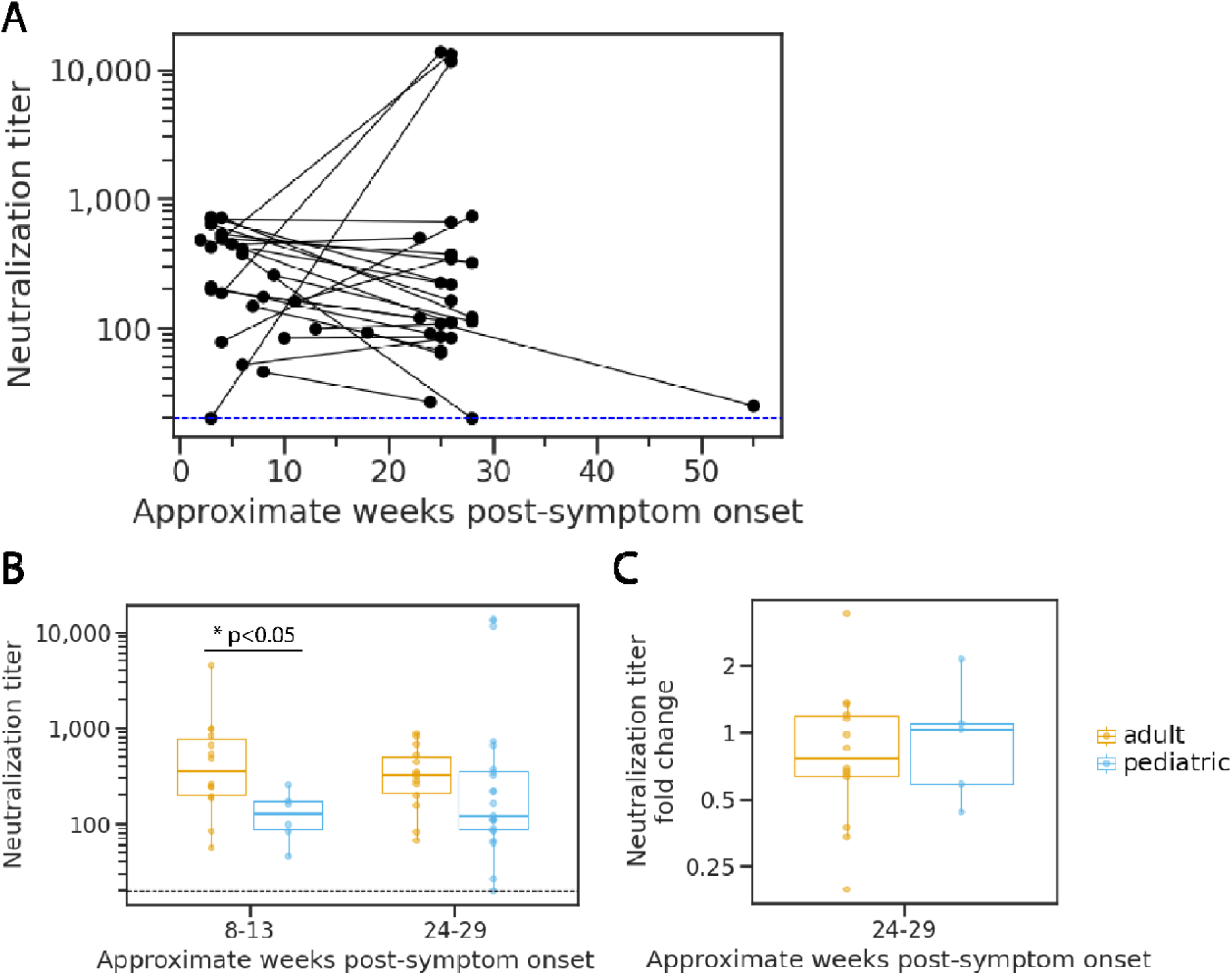
Neutralization potency kinetics in children compared to adults. **A**) Aggregated trajectories of pediatric neutralization titers (NT_50_) longitudinally with lines connecting specimens from the same individual for the 25 pediatric participants without underlying immunosuppression, receipt of multiple blood transfusions, or MIS-C. **B**) Comparison of adult and pediatric neutralization titers collected within the time periods 8 to 13 weeks (adults N = 14; children N = 7) and 24 to 29 weeks (adults N = 14; children N = 22) for the participants without underlying immunosuppression, receipt of multiple blood transfusions, or MIS-C. **C**) Analysis of fold change in neutralization titers at 24 to 29 weeks (adults N = 14; children N = 6) relative to titers at 8 to 13 weeks for adults and children without underlying immunosuppression, receipt of multiple blood transfusions, or MIS-C. Significance determined by Mann Whitney test.

For further clinical and laboratory data on children with underlying immunocompromising conditions, multiple blood transfusions, or MIS-C, please refer to Figures 2B & C. Three children with specimens at 52 weeks had detectable neutralizing antibodies (**Figure 2A, B, & C**). Of note, one child (C26) with blood collected at 52 weeks reported a febrile illness, with negative SARS-CoV-2 RT-PCR, between the 24- and 52-week specimen collection (**Figure 2C**).

### Comparison of neutralization dynamics in immunocompetent children and older adults

We next compared neutralization titers and their longitudinal dynamics in children and adults. To accomplish this, we measured plasma neutralizing antibody levels from adults over a 24-week period. Neutralization titers for specimens collected at 8- to 13-weeks post-symptom onset (first collection period) were previously reported using the same spike pseudotyped lentivirus neutralization assay but without the D614G spike mutation (3). Here, we repeated the neutralization assays using spike pseudotyped lentivirus encoding D614G as well as performing neutralization assays for the first time on specimens collected between 24 and 29 weeks (second collection period). Neutralization titers had a geometric mean of 385 (range: 56 - 4,487) and 302 (range: 67 – 880) at the first and second collection period, respectively (**Supplemental figure 2**). Of the 14 participants in our adult cohort, only one demonstrated a greater than 4-fold decrease in neutralization titer over the observation period, and no adults showed an increase greater than 4-fold. There were no adults for whom neutralization titers fell below the limit of detection during the timeframe tested.

For comparison of neutralization titers between the children and adults including older adults, we restricted our analysis to only specimens collected in the same timeframe for both cohorts, as well as only including children without immunocompromising conditions, those who did not receive multiple blood transfusions, and those without MIS-C. In this sub-analysis, we found that children had significantly lower neutralization potency (geometric mean titer [GMT] = 118, range: 46-256, N=7, *p*<0.05) than adults (GMT = 385, range: 56-4,487, N=14) during the first collection period, but titers were not significantly different between age groups by the second collection period (children: GMT= 244, range: 27-13,694, N=22; adults: GMT = 302, range: 67-880, N=14; *p* = 0.23) (**Figure 3B**). If the four children with neutralization titers that increased by greater than 4-fold are excluded, the children’s GMT for the second collection period is 2.46-fold lower than the adults’ (123 compared to 302 in children and adults, respectively). We calculated the fold change in titers for each individual measured at the first collection period relative to those measured for the same individual during the second time period. Fold change analysis was limited to 5 children with specimens collected at both first and second collection period; no difference in the fold change between children (geometric mean fold decrease = 1.12, N=6,) and adults (geometric mean fold decrease = 1.28, N=14) was detectable *(p* = 0.893). (**Figure 3C**).

### Anti-nucleocapsid antibody dynamics over time in children

Anti-N antibody levels were determined for all pediatric specimens (**Figure 4A, B, & C**). Among the 25 children without immunocompromising conditions, multiple blood transfusions, or MIS-C, 23 and 14 had detectable anti-N antibodies at the first and second collection periods, respectively; 2 children with confirmed infection by RT-PCR (C1 and C32) did not have detected anti-N antibodies at either timepoint. Anti-N antibody levels dropped considerably from a geometric mean index of 3.7 to 1.3 over 24 weeks. Eighteen of the 23 children, who were positive for anti-N antibodies at the first collection period, exhibited a decrease in index values of greater than 2-fold, and an additional five changed less than 2-fold. No children showed an increase in anti-N antibodies. In totality, the children without immunocompromising conditions showed very similar declining trends in anti-N antibody levels across time (**Figure 5A**). Of the children with a positive index at 4 weeks, values ranged from 1.9 to 8.0 and from undetectable to 7.3 by the first and second collection periods, respectively.

**Figure 4.**
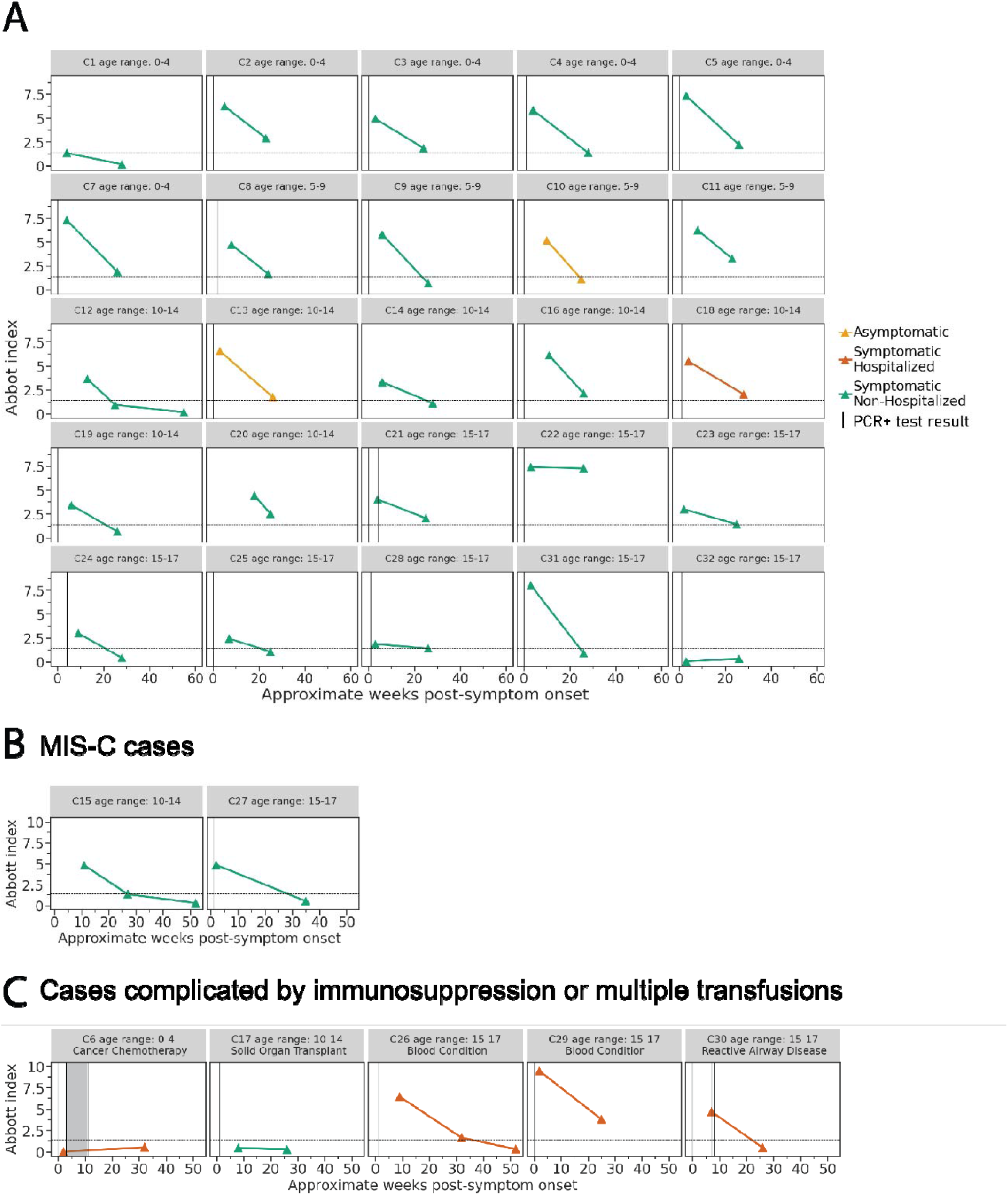
Anti-nucleocapsid antibody binding in children over time. Anti-N antibody titers in **A)** 25 children with confirmed SARS-CoV-2 infection, **B)** children who developed MIS-C following acute infection, and **C)** cases complicated by immunosuppression or multiple blood transfusions in 5 children with confirmed SARS-CoV-2 infection followed prospectively over time shown as weeks. Vertical lines represent the week of positive RT-PCR test result(s), and shaded areas indicate weeks with consecutive positive RT-PCR test results. Colors show disease severity during acute infection. Dotted horizontal lines indicate the limit of detection for the SARS-CoV-2 IgG assay (1.40).

**Figure 5.**
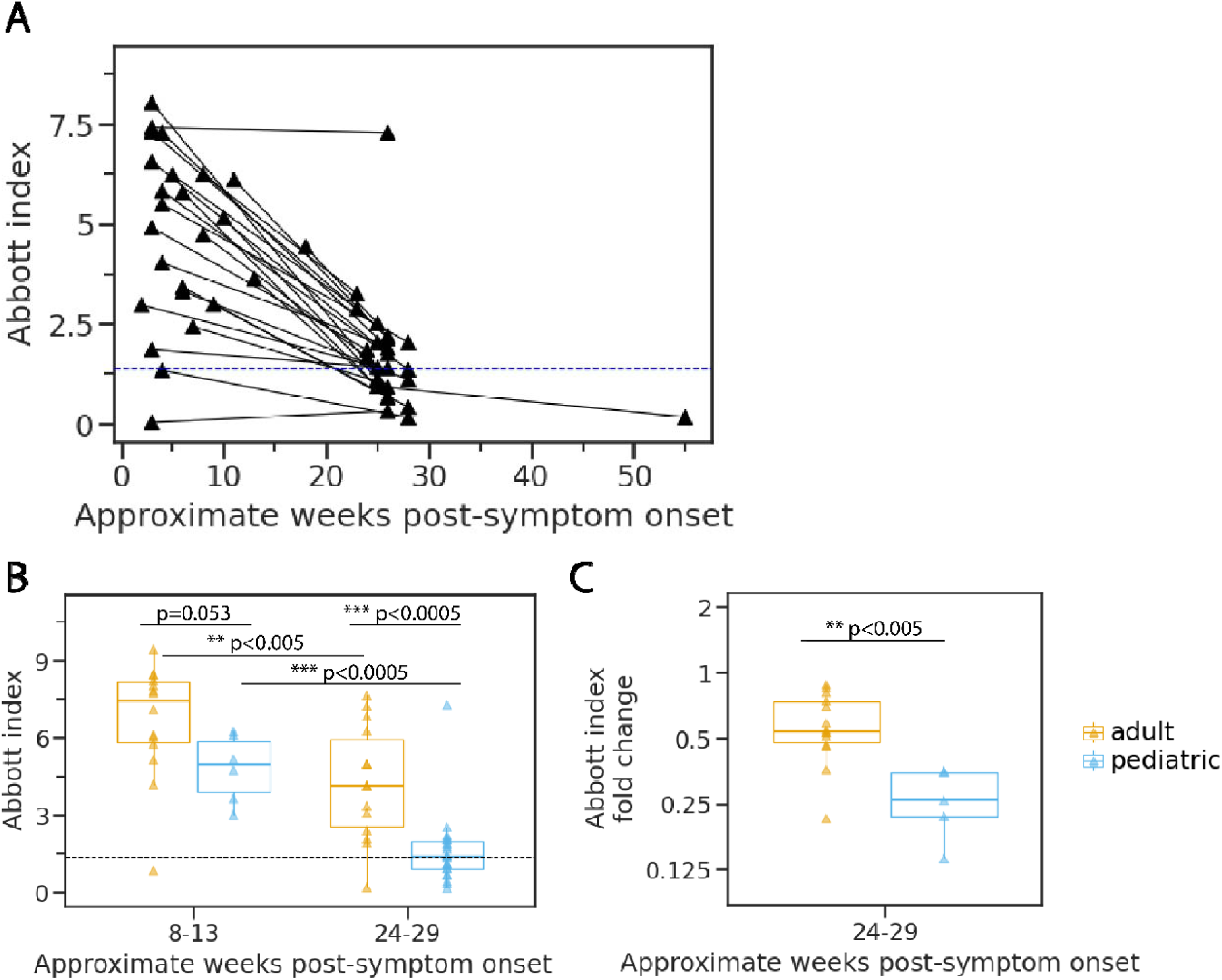
Change in nucleocapsid-binding antibody levels longitudinally in children and adults. **A**) Aggregated index values for children without immunocompromising conditions over one-year post-symptom onset with lines connecting specimens from the same individual. **B**) Comparison of index values between pediatric and adult cohorts restricted to the same time periods of collection. **C**) Change in index values at 24 to 29 weeks relative to specimens collected at 8 to 13 weeks for children and adults with specimens collected within both timeframes. Significance determined by Mann Whitney test. Dotted lines indicate the limit of detection for the SARS-CoV-2 IgG assay (1.40).

For anti-N antibody levels and clinical information for the children with underlying immunocompromising conditions, multiple blood transfusions, or MIS-C refer to Figure 4B & C. The antibody dynamics out to 52-weeks post-symptom onset were measured for three children all of whom had levels below the limit of detection by this later time period (**Figure 4A, B, &C**).

### Comparison of pediatric and adult anti-nucleocapsid antibody dynamics

Next, we compared anti-N antibody dynamics in children and adults. We first measured anti-N antibody levels for all adult specimens in our cohort (**Supplemental figure 3**). Overall, geometric mean values in adults fell from 6.0 to 3.3 between the first and second collection period, respectively. One adult (A12) had values below the limit of detection at both 8- and 24-weeks post-symptom onset. Of the adults with a positive index at 8 weeks, values ranged from 4.2 to 9.4 and from 1.9 to 7.7 by the first and second collection period, respectively. No adults with positive index values at the first timepoint fell below the limit of detection by the later timepoint. This is in stark contrast to the pediatric cohort where many fell below detectable levels over the course of the study. Furthermore, only 3 adults showed a greater than 2-fold decrease in index values.

Compared to the pediatric cohort, adults had higher anti-N antibody levels at both timepoints measured although not quite reaching statistical significance at 8-13 weeks (children: GMT = 4.7, range: 3.0-6.2; adults: GMT = 6.0, range: 0.8-9.4; p=0.053) (**Figure 5B**). The difference between adult and child index values was greatest at the later 24- to 29-week timepoint (children: GMT = 1.2, range: 0.2-7.3; adults: GMT = 3.3, range: 0.2-7.7; p<0.0005) suggesting that anti-N antibodies may wane faster in children than adults. To test this, we compared the fold change between the first and second collection periods in children and in adults. We found a greater decrease for the pediatric cohort (geometric mean decrease of 4-fold) demonstrating that these children lost N antibody binding at a faster rate than the adult cohort (geometric mean decrease of 1.8-fold) (**Figure 5C**).

## Discussion

In this study, we describe the kinetics of serum antibodies over time in children after infection with SARS-CoV-2. In our convenience samples of unvaccinated children and adults with confirmed or presumed SARS-CoV-2 infection, we found that pediatric serum neutralizing titers were maintained over 24 weeks while anti-N-binding antibodies waned quickly. Importantly, neutralizing antibody titers were highly variable among individual children as has been previously observed in adults (1, 3, 6, 8, 10, 11, 23, 24, 45). Other studies have demonstrated that greater disease severity and higher viral load are associated with higher antibody levels in adults (3, 10, 46). The limited number of asymptomatic, hospitalized, and MIS-C cases in our cohort prevented analysis of the role that disease severity may play in this variability. While further investigation is needed, the wide range of neutralization titers and anti-N antibody levels observed in our group of 22 immunocompetent, non-MIS-C presenting children, who were symptomatic but not hospitalized, suggests that disease severity may not entirely explain the observed heterogeneity.

There are several reasons why antibody responses to SARS-CoV-2 infection could be different in children compared to adults, including disease typically being less severe in children (21, 47–51) as well as immune senescence and greater burden of comorbidities in older adults (52–58).

Further, primary infections with respiratory pathogens tend to occur early in life leaving uncertainty about how antibody responses to primary infection may differ with age. Additionally, children are susceptible to life threatening MIS-C following infection, and it remains unclear if and/or how the immune response following infection may impact development of such sequelae.

Interestingly, only a modest and non-significant decrease in neutralizing antibody level was detected for pediatric specimens collected out to six months. A similar persistence in neutralization potency was also observed in the adult cohort, suggesting that there might be long term maintenance of neutralizing antibodies regardless of age following SARS-CoV-2 infection. This finding is in line with several other bodies of work demonstrating the persistence of neutralizing antibodies over many months (9, 26, 59–61). We did, however, detect lower levels of neutralization in children’s serum compared to adults early after infection. This finding is perhaps surprising given recent work, in the context of vaccination, showing that older adults, similar to the age group of adults reported here, develop lower neutralizing titers than younger adults (62). Antibody dynamics across ages may be different between infection and vaccination, and other factors such as specimen collection time or disease severity could also contribute the difference between this study and ours. Interestingly, by 24 weeks, a difference in neutralization titers between children and adults was no longer detectable. This leveling of neutralization titers over time has also been observed for some (3) but not all (11) studies of adults who have disease of different severity: adults with severe disease have higher initial titers at early, but not later, timepoints (3). Overall, the neutralizing antibody kinetics that we observe for children are similar to adults with mild infections (3, 14). A previous study corroborates our findings of lower pediatric neutralization titers early after infection by measuring neutralization titers in children and adults out to 60 days (24), and another study looking at only hospitalized children and adults reported the same (63). However, one study (26) found that younger children had higher titers than older children and adults. Differences in study population and sampling timepoints could explain these differences.

The most striking difference in SARS-CoV-2 antibody levels between children and adults was seen for anti-N antibodies. Although not statistically significant, children tended to have lower levels than adults early after infection and a significantly lower level after six months. Lower anti-N antibody levels in children than adults have been reported in another study as well (24). Those authors speculated that, since nucleocapsid protein is disseminated during infection through the lysis of infected cells, children may experience lower levels of N antigen expression due to their reduced duration of illness and potentially lower levels of viral replication (24).

Alternatively, the cumulative lifetime exposure to betacoronavirus infections in adults may repeatedly boost antibodies to the more conserved nucleocapsid proteins that are cross-reactive to SARS-CoV-2, as has been observed for conserved influenza proteins (64). It is important to note that several studies have found that the SARS-CoV-2 IgG assay used for this study decreases in sensitivity over time faster than in other assays (13, 23, 40–44). In addition, the SARS-CoV-2 IgG assay only has emergency use authorization for qualitative assessment of antibodies and not quantitative.

Limitations of our study include small sample size, a limited number of children with follow-up at 52-weeks, and differences in the sex distribution between the pediatric and adult cohorts. Follow-up is ongoing with children who had not yet reached 52-weeks post-symptom onset at the time of this analysis. Furthermore, blood volume obtained from younger children is limited and therefore the number of assays utilized was also limited. The adult comparative specimens were obtained from the same geographic location and analyzed in the same laboratory, although not necessarily collected from the same families or at the same time. The adult specimens were also plasma, whereas the pediatric specimens were serum, and the differences in collection and storage of these could possibly result in slight differences in antibody concentrations.

Additionally, the adults in this study were a convenience sample of a broader study, and approximately half were older adults, over 65 years of age, meaning that the data presented here may not be representative of all adults across wider age ranges. Likewise, our pediatric cohort was also a convenience sample and may also not be representative of the broader population. Furthermore, unlike the pediatric cohort, adults were only enrolled following RT-PCR confirmed infection without enrollment based on household RT-PCR positive contacts. Of note, both children and adult cohorts were enrolled prior to the widespread introduction of the SARS-CoV-2 Delta variant.

Overall, our results suggest that although neutralizing antibody responses to SARS-CoV-2 are broadly similar between adults and children, anti-N antibodies are elicited at lower levels in children than adults. These results contribute to our knowledge of pediatric immune responses to SARS-CoV-2 over time, and the data on the longevity of neutralizing antibodies may prove valuable for comparison investigations of immunity induced by vaccines in children.

## Data Availability

All data produced are contained in the manscript or its supplementary material.

## Acknowledgements

The authors would like to acknowledge Andrea Loes for her supportive role in this study. This work was supported by CDC BAA75D301-20-R-67897 and also by the NIAID/NIH R01AI141707 and R01AI127893 grant to J.D.B. The findings and conclusions in this report are those of the author(s) and do not necessarily represent the official position of the Centers for Disease Control and Prevention (CDC). In addition, J.D.B. is an Investigator of the Howard Hughes Medical Institute. This material is based upon work supported by the National Science Foundation Graduate Research Fellowship Program under Grant No. DGE-1762114 to L.E.G. Any opinions, findings, and conclusions or recommendations expressed in this material are those of the author(s) and do not necessarily reflect the views of the National Science Foundation. The funders had no role in study design, data collection, or the decision to submit the work for publication.

J.D.B consults for Moderna, Oncorus, and Flagship Labs 77. J.D.B. is an inventor on Fred Hutch licensed patents related to deep mutational scanning of viral proteins. J.A.E. consults for AstraZeneca, Sanofi Pasteur, Meissa Vaccines, Teva Pharmaceuticals, and receives research support from AstraZeneca, Merck, Novavax, and Pfizer. H.Y.C. receives research support from Gates Ventures, NIH, CDC, DARPA, Sanofi-Pasteur, and Cepheid and serves on the advisory boards for Merck, Pfizer, Ellume, and the Gates Foundation. All other authors report no additional competing interests.

Pediatric study design, patient enrollment and data acquisition, and specimen collection were completed by J.A.E., L.K., K.L., J.Y., and S.S. Adults study design, patient enrollment and data acquisition, and specimen collection was completed by H.Y.C. and L.W. Neutralization assays, data analysis, and manuscript writing were performed by L.E.G. All work was performed under the direction of J.D.B. and J.A.E. SARS-CoV-2 IgG assay were performed by J.D. Manuscript editing and review was completed by J.D.B, J.A.E., H.Y.C., L.K., K.L., S.N., S.S., K.P., M.B.H, and C.M.M.

## Supplemental figures

**Supplemental Table 1.**
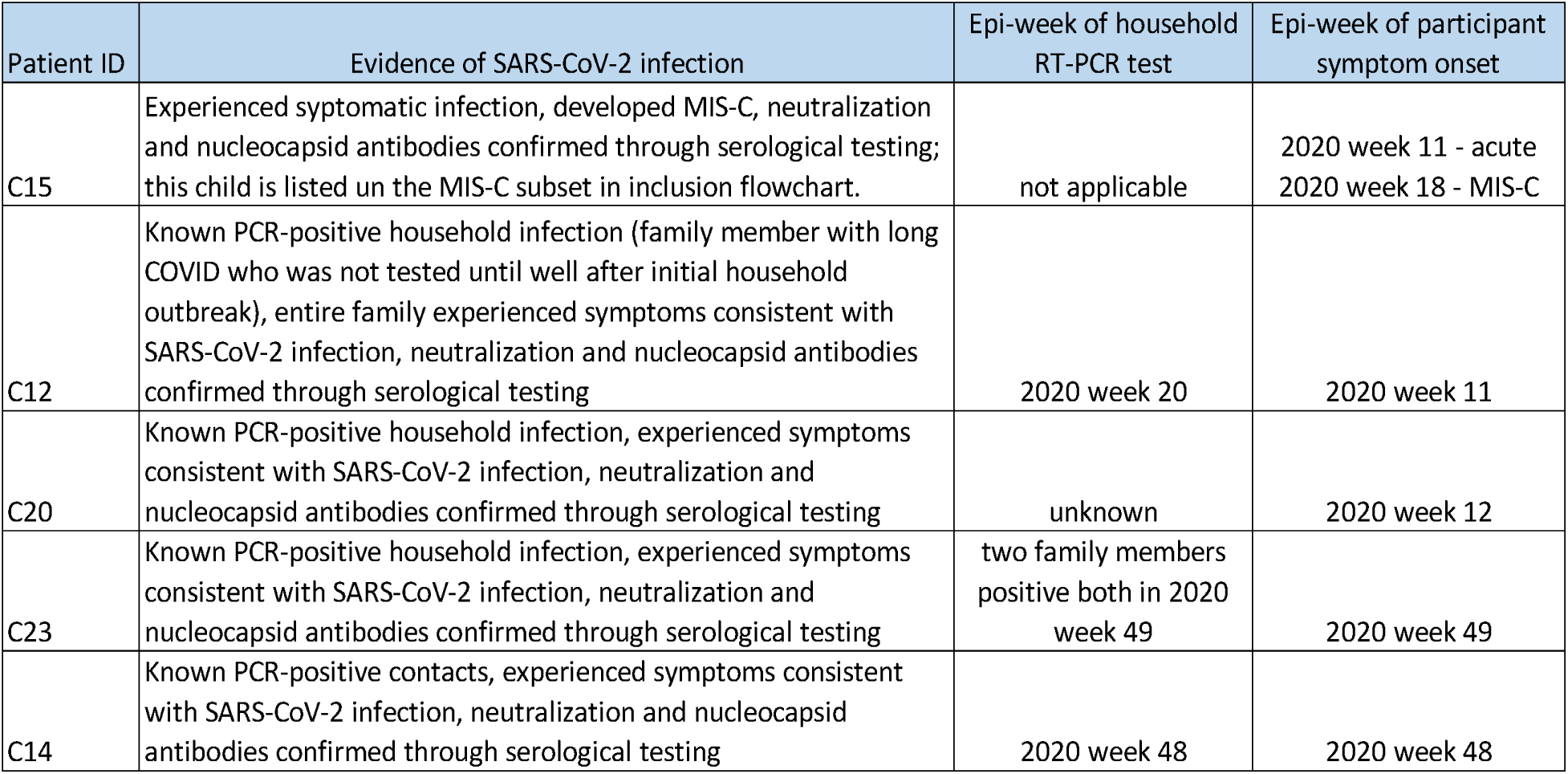
Evidence of SARS-CoV-2 infection among patients without a confirmed SARS-CoV-2 RT-PCR.

| Patient ID | Evidence of SARS-CoV-2 infection | Epi-week of household RT-PCR test | Epi-week of participant symptom onset |
| --- | --- | --- | --- |
| C15 | Experienced symptomatic infection, developed MIS-C, neutralization and nucleocapsid antibodies confirmed through serological testing; this child is listed on the MIS-C subset in inclusion flowchart. | not applicable | 2020 week 11 - acute<br>2020 week 18 - MIS-C |
| C12 | Known PCR-positive household infection (family member with long COVID who was not tested until well after initial household outbreak), entire family experienced symptoms consistent with SARS-CoV-2 infection, neutralization and nucleocapsid antibodies confirmed through serological testing | 2020 week 20 | 2020 week 11 |
| C20 | Known PCR-positive household infection, experienced symptoms consistent with SARS-CoV-2 infection, neutralization and nucleocapsid antibodies confirmed through serological testing | unknown | 2020 week 12 |
| C23 | Known PCR-positive household infection, experienced symptoms consistent with SARS-CoV-2 infection, neutralization and nucleocapsid antibodies confirmed through serological testing | two family members positive both in 2020 week 49 | 2020 week 49 |
| C14 | Known PCR-positive contacts, experienced symptoms consistent with SARS-CoV-2 infection, neutralization and nucleocapsid antibodies confirmed through serological testing | 2020 week 48 | 2020 week 48 |

**Supplemental figure 1.**
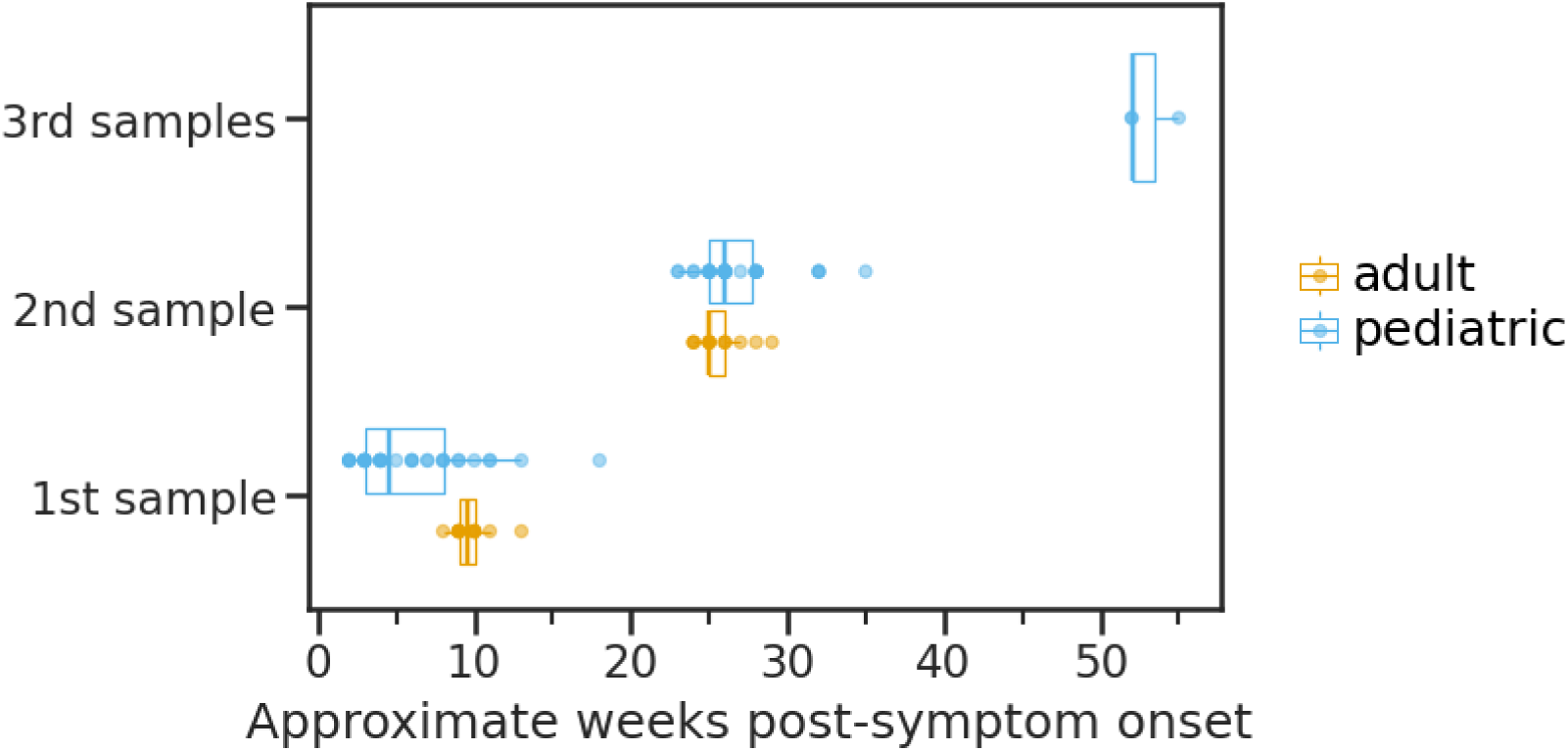
Distribution of specimen collections in children and adults.

**Supplemental figure 2.**
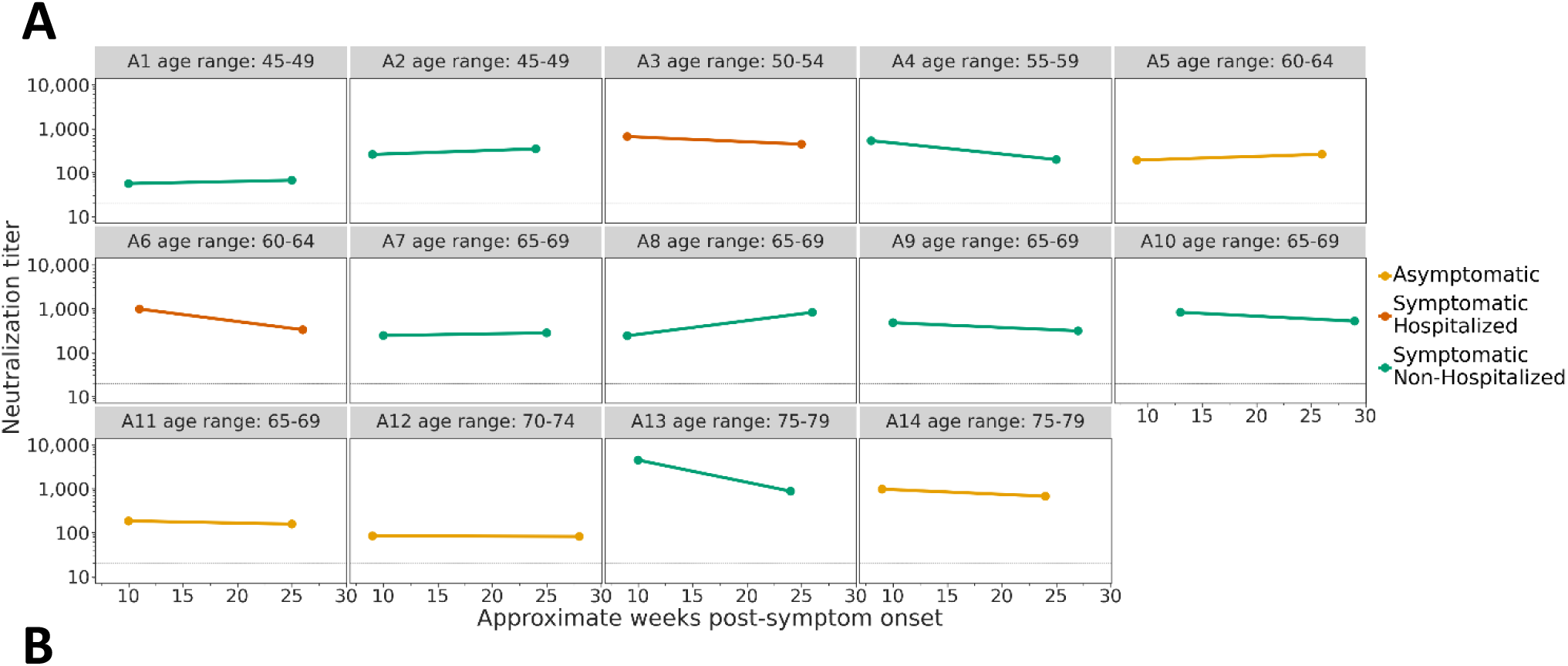
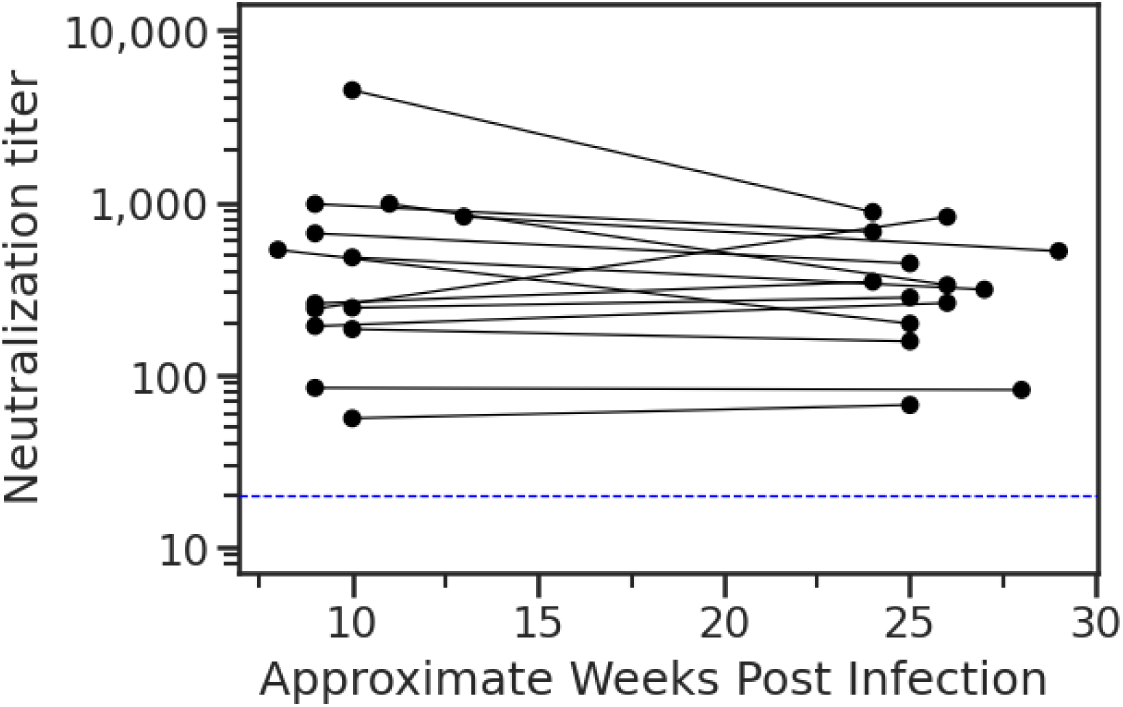
Neutralization titers in adults over time. **A**) Neutralizing antibody titers in 14 adults with confirmed SARS-CoV-2 infection followed prospectively over time shown as weeks post-symptom onset, x axis. **B**) Aggregated neutralization titers for all adults. Dotted horizontal lines indicate the limit of detection (20).

**Supplemental figure 3.**
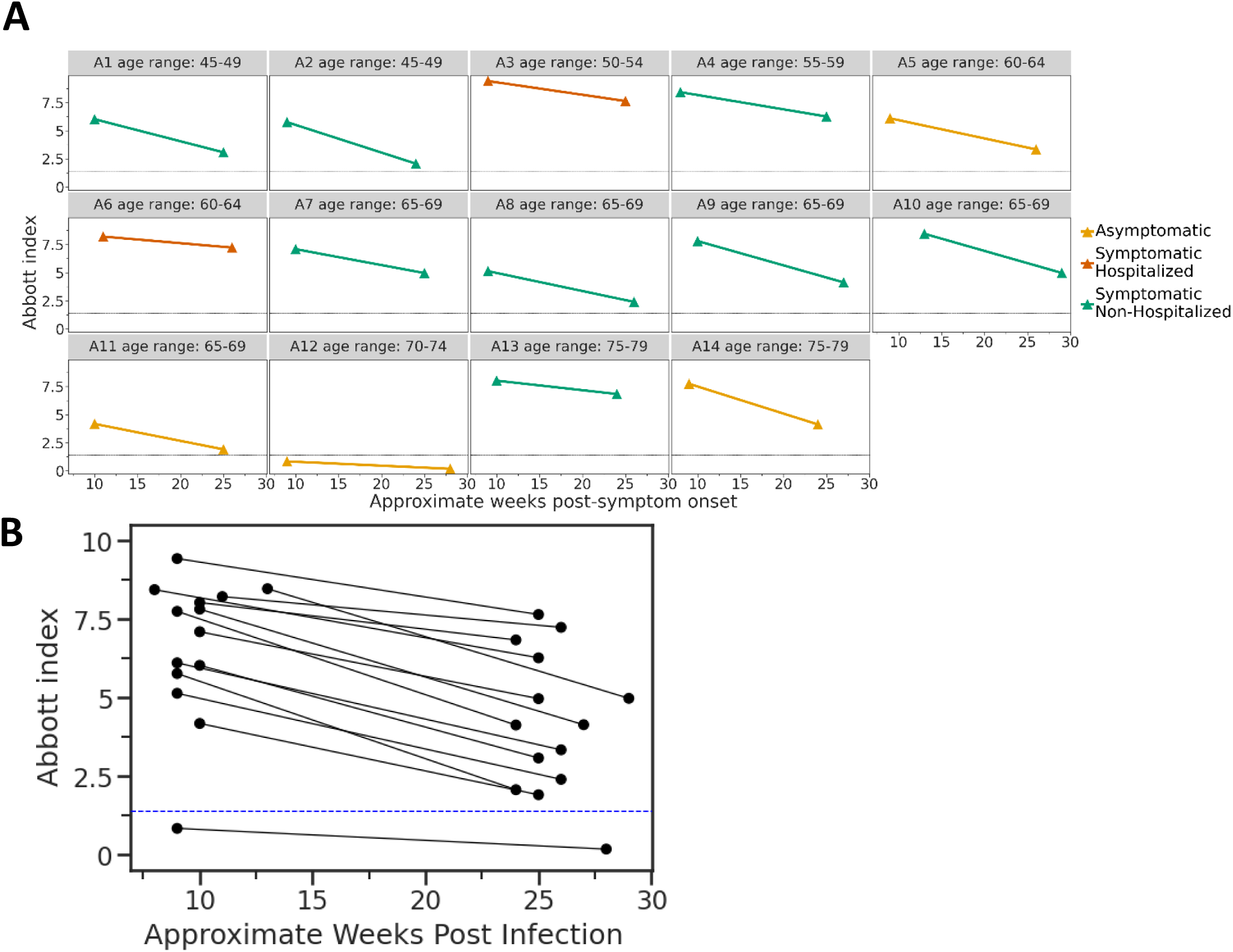
Nucleocapsid-binding antibody levels in adults over time. **A**) The SARS-CoV-2 IgG assay was used to determine SARS-CoV-2 nucleocapsid-binding antibody in 14 adults followed prospectively over time shown as weeks post-symptom onset, x axis. **B**) Aggregated index values for all adults. Dotted horizontal lines indicate the limit of detection for the SARS-CoV-2 IgG assay (1.40).

**Supplemental Table 2.** Naming of adults across publications.

| Naming in Crawford et al. 2020 (3) | Naming in the present study |
| --- | --- |
| PID 13 | A3 |
| PID 3C | A1 |
| PID 4C | A2 |
| PID 6C | A6 |
| PID 7C | A7 |
| PID 11C | A4 |
| PID 12C | A10 |
| PID 22C | A9 |
| PID 23C | A8 |
| PID 24C | A13 |
| PID 103C | A12 |
| PID 113C | A14 |
| PID 117C | A11 |
| PID 200C | A5 |

